# Does combination treatment with progesterone and vaginal cervical cerclage improve pregnancy outcomes in women at high risk of preterm birth? A retrospective, secondary analysis of the C-STICH suture thread for cerclage trial data

**DOI:** 10.1101/2024.12.23.24319536

**Authors:** V Hodgetts Morton, RK Morris, P Toozs-Hobson, L Middleton, N Pilarski, L Bell, M Hogg, R Man, F Israfil-Bayli, A Shennan, N Simpson, C Lees, CA Moakes

## Abstract

**Background:** Vaginal cervical cerclage and progesterone are established treatments for prevention of pregnancy loss and prematurity. There is limited data to assess the effect of these treatments in combination.

**Objective:** To investigate the association between progesterone and no progesterone treatment on pregnancy outcomes in women at high risk of preterm birth who have received a vaginal cervical cerclage.

**Study Design:** This is a secondary analysis of women recruited to the CSTICH trial, which recruited in 75 obstetric units in the UK. The primary outcome was pregnancy loss, defined as miscarriage and perinatal mortality, including any stillbirth or neonatal death in the first week of life. Secondary maternal outcomes included miscarriage and previable neonatal death; stillbirth; gestational age at delivery; preterm pre labour rupture of membranes and sepsis. Secondary neonatal outcomes included early/late neonatal death and sepsis. For each outcome, regression models were fitted adjusting for pre-specified prognostic variables.

**Results:** 1943 women had a vaginal cerclage placed, with available progesterone data. From the 2048 women recruited to CSTICH; 843 (43%) women received progesterone and 1109 (57%) did not receive progesterone. Pregnancy loss occurred in 49 (5.9%) of 832 women who received progesterone and 91 (8.3%) of 1103 women who did not receive progesterone (adjusted risk ratio 0.70, 95% confidence interval (CI): 0.50 to 0.99; adjusted risk difference - 0.02, 95% CI: −0.04 to −0.001).

**Conclusion:** There appears to be an association with progesterone use in women who receive a vaginal cervical cerclage and a reduction in pregnancy loss. This combination therapy may be an important opportunity to further reduce the risk of pregnancy loss in this high risk cohort.

## Introduction

Second trimester miscarriage and preterm birth is a multifactorial condition affecting approximately 10% of pregnancies. Second trimester miscarriage and preterm birth are a continuum of a conditions with overlapping aetiology.(1) The prematurity syndrome does not have a singular cause, with cervical weakness, inflammation and infection, stress, uterine overdistension and placental/vascular disorders all contributing to a poorly understood process that potentially prematurely initiates the labour process.(1)

Vaginal cervical cerclage and vaginal progesterone are both established treatments to reduce the risk of second trimester miscarriage and preterm birth.(2, 3) The evidence for progesterone and vaginal cervical cerclage have been examined primarily as single interventions in the same population of women at high risk of pregnancy loss.(4, 5) It is therefore reasonable to consider whether these treatments have additional benefit in combination, working synergistically to improve pregnancy outcomes. The evidence for the use of vaginal progesterone and cervical cerclage in combination is limited and there are no randomised controlled trials designed specifically to answer this question. Roman et al. described the potential benefit of rescue vaginal progesterone in women who had continued cervical shortening despite the placement of a vaginal cervical cerclage, demonstrating a potential prolongation of pregnancy (36.36 weeks vs. 32.63 weeks; p = 0.0036).(6) This was a small study consisting of 66 women but highlights the need to consider whether these treatments are synergistic.

C-STICH was a completed randomised controlled trial of type of suture thread (monofilament vs braided) in women undergoing a vaginal cervical cerclage.(7) C-STICH recruited 2048 eligible women where the clinical decision for a vaginal cervical cerclage had already been made. Women were then randomised prior to the cervical cerclage surgery to either a monofilament or braided suture thread. The trial was pragmatic in that the operative technique/adjuncts, decision to use progesterone and clinical management were all at the discretion of the local recruiting clinical team. Within C-STICH there was no difference in the primary outcome of pregnancy loss (defined as miscarriage and perinatal mortality, including any stillbirth or neonatal death in the first week of life). (8) The data collected within C-STICH has provided the opportunity to examine the effect of receiving progesterone on pregnancy loss, prematurity and maternal and neonatal outcomes, in women who undergo a vaginal cervical cerclage.

### Aims and objectives

The overarching aim of this analysis was to examine the effect of combination treatment of progesterone and vaginal cervical cerclage, compared with vaginal cervical cerclage alone on pregnancy outcomes in women at high risk of preterm birth.

The primary objective was to investigate whether vaginal progesterone reduces the risk of pregnancy loss, in women undergoing a vaginal cervical cerclage due to an increased risk of preterm birth. Secondary objectives were to assess the characteristics of women (including indication for cerclage) who did or did not receive progesterone and to assess whether in women undergoing a vaginal cervical cerclage due to an increased risk of preterm birth, vaginal progesterone improves other pregnancy and neonatal outcomes.

## Methods

This is a secondary analysis of participants in the CSTICH trial which recruited in 75 obstetric units in the UK and was published previously (5, 6).

### Patients

Women were considered eligible for the CSTICH trial if they had an indication for a vaginal cervical cerclage, were aged 18 years or older and had a singleton pregnancy. Indication for cervical cerclage was defined as either a history of three or more previous mid-term losses or premature births (≤28 weeks), insertion of cervical sutures in previous pregnancies, a history of mid-trimester loss or premature birth, with a shortened cervix (≤25 mm) in the current pregnancy, or clinician concern for risk of preterm birth either due to history or the results of an ultrasound scan. Women were ineligible if they required an emergency or rescue cerclage; needed immediate insertion of a suture; had membranes that had ruptured or were visible, or required a cerclage which was to be placed by any route other than vaginally (e.g., via an abdominal route). All participants provided written informed consent. Additional inclusion criteria for this secondary analysis were women who had available progesterone data and had a vaginal cervical cerclage successfully placed.

### Intervention group

Combination treatment of progesterone and vaginal cervical cerclage.

### Comparator group

Vaginal cervical cerclage alone.

### Outcomes

The primary outcome is pregnancy loss rate defined as miscarriage and perinatal mortality, including any stillbirth or neonatal death in the first week of life. A key secondary outcome was time from conception to pregnancy end. Secondary outcomes included the following maternal and neonatal outcomes:

Maternal

− Miscarriage & previable neonatal death (defined as delivery <24 weeks).
− Stillbirth (defined as intrauterine death ≥24 weeks).
− Gestational age at delivery (in live births ≥24 weeks).
− Gestational age <28/<32/<37 weeks at delivery (in live births ≥24 weeks).
− Preterm pre labour rupture of membranes (PPROM).
− Sepsis (at any time in pregnancy and until 7 days postnatal).

Neonatal

− Early neonatal death (defined as a death within 7 days after delivery, in live births ≥24 weeks).
− Late neonatal death (defined as a death beyond 7 days and before 28 days after delivery, in live births ≥24 weeks).
− Sepsis (clinically diagnosed/proven) (in live births ≥24 weeks).

### Sample size

There were 1943 participants within CSTICH who had a vaginal cerclage successfully placed where progesterone data was available. Whilst this sample size was not specifically tailored to this secondary analysis, this size of sample would be sufficient to detect a reduction in pregnancy loss from 9.3% in the combination treatment of progesterone and vaginal cervical cerclage group, to 5.7% in the vaginal cervical cerclage only group, with 85% power.

### Statistical analysis

A statistical analysis plan was pre-specified before analysis. All estimates of differences between groups were analysed using regression models presented with two-sided 95% confidence intervals (CIs) adjusted for primary indication for cerclage (a history of three or more previous midterm losses or premature births (≤28 weeks)/Insertion of cervical sutures in previous pregnancies/A history of mid trimester loss or premature birth with a (current) shortened (≤25mm) cervix/Women whom clinicians deem to be at risk of preterm birth either by history or the results of an ultrasound scan); number of previous mid trimester losses; number of previous pre-term births (<34 weeks); any previous cervical surgery (yes/no/unknown); cerclage technique involved bladder dissection (yes/no); suture type received (monofilament/braided); ethnicity (white/black/asian/mixed/other/declined to provided information) and maternal age at cerclage placement (where possible). Time from conception to live birth and gestational age at delivery were further adjusted for gestational age at cerclage placement. If full covariate adjustment was not possible (e.g. the model did not converge), covariates were removed sequentially in the reverse order to those listed above. The variables for adjustment within this analysis are consistent with the randomisation stratification within the primary randomised controlled trial, determined by an expert group to have a significant effect on the overall risk of pregnancy loss.

Baseline data were summarised descriptively and included those listed above as adjustment covariates. Tests of statistical significance were undertaken using t-tests or Wilcoxon rank sum tests for continuous variables (depending on the distribution of the data) and chi-square or Fishers exact tests for categorical variables.

The primary outcome was summarised using frequencies and percentages. A mixed effects log-binomial model was used to generate an adjusted risk ratio (RR) (and 95% CI). An adjusted risk difference (RD) (and 95% CI) was also presented (using an identity link function). Binary secondary outcomes (miscarriage & pre-viable neonatal death, stillbirth, gestational age <28/<32/<37 weeks, PPROM, early neonatal death and late neonatal death) were analysed as per the primary outcome. Continuous secondary outcome measures (gestational age at delivery) were summarised using means and standard deviations alongside an adjusted mean difference (with 95% CI) estimated using a linear regression model.

Time to event data (time from conception to pregnancy end) were considered in a competing risk framework. This was to account for the different outcomes of pregnancy end. A cumulative incidence function was used to estimate the probability of a live birth (surviving at least seven days) over time, accounting for the competing event of pregnancy loss. This method is favoured over Kaplan-Meier methods which may overestimate this probability. A Fine-Gray model was used to estimate a subdistribution adjusted hazard ratio (HR) (and 95% CI) directly from the cumulative incidence function. The subdistribution HR estimates the overall incidence of live birth (i.e. presence of live birth over time). In addition, a Cox Proportional Hazard model was fitted and applied to the cause-specific hazard function and used to generate a cause-specific adjusted HR (and 95% CI) which estimates the rate in which live births occur (9, 10).

Since the proportion of participants with missing data for the primary outcome was <1% (N=8), no supporting analyses to assess the impact of missing data were conducted (i.e. complete case analyses only).

We conducted prespecified subgroup analyses (limited to the primary outcome measure only) for number of previous mid trimester losses (<3/≥3) and number of previous pre-term births (<34 weeks) (0/≥1). The effects of these subgroups were examined by adding the subgroup by treatment group interaction parameters to the regression model. P-values from the tests for statistical heterogeneity were presented with the effect estimate and estimates of uncertainty within each subgroup. Additionally, ratios were provided to quantify the difference between the treatment effects estimated within each subgroup.

All analyses were performed in SAS (version 9.4) or Stata (version 18.0).

## Results

Of the 2048 women randomised to C-STICH, 1998 women had a cerclage placed, with progesterone data available in 1943 women. Of these, 834 (43%) received progesterone treatment in combination with their vaginal cervical cerclage. Progesterone was commenced greater than 7 days prior to cerclage placement in 191 (25%) of cases, commenced within a week of cerclage placement in 298 (39%) of cases and greater than 7 days after cerclage placement in 277 (36%) of cases (detailed in table 1).

**Table 1:** Progesterone information:

|  | Number randomised to the C-STICH trial (N=2048*) |
| --- | --- |
| Cerclage placed-N (%) |  |
| Yes | 1998 (98) |
| No | 45 (2) |
| Missing | 5 |
| Progesterone use<sup>1</sup>-N (%) |  |
| Yes | 834 (43) |
| No | 1109 (57) |
| Missing | 55 |
| Gestational age progesterone received (weeks)<sup>2</sup> |  |
| Median [IQR, N] | 16.0 [13.6-19.1, 766] |
| Minimum-Maximum | 0-34.6 |
| Missing | 68 |
| Time from cerclage placement to progesterone start (days)<sup>2</sup>-N (%) |  |
| ≥7 days prior to cerclage placement | 191 (25) |
| <7 days prior to cerclage placement or <7 days after cerclage placement | 298 (39) |
| ≥7 days after cerclage placement | 277 (36) |
| Median [IQR, N] | 1.0 [-6-13, 766] |
| Minimum-Maximum | -133-151 |
| Missing | 68 |
| Progesterone type<sup>2,3</sup>-N (%) |  |
| Cyclogest 200mg PV daily | 239 (29) |
| Cyclogest 400mg PV daily | 429 (53) |
| 17 alpha-hydroxyprogesterone 250mg IM weekly | 117 (14) |
| Other | 71 (9) |
| Missing | 22 |

\*Excluding the woman who was randomised in error as no data was collected.
<sup>1</sup>In women who had a cerclage placed.
<sup>2</sup>In women who had a cerclage place and received progesterone.
<sup>3</sup>Progesterone types are not mutually exclusive.

The demographics of the included participants are detailed in table 2. There were notable differences between those that did not receive additional progesterone treatment and those who did. Women with no previous live pre-term births were more likely to receive combined treatment as were those who underwent a cervical cerclage with bladder dissection (Shirodkar technique, 17% in those who did not receive progesterone vs. 34% in those who received progesterone, p<0.01), were non-black ethnicity (78% in those who did not receive progesterone vs. 85% in those who received progesterone, p=0.01) and those whose indication for cerclage was due to risk of preterm birth through history or ultrasound (58% in those who did not receive progesterone vs. 63% in those who received progesterone, p=0.03). The median gestational age at cerclage placement was 16.3 weeks in those who did not receive progesterone and 15.9 weeks in those who received progesterone (p=0.02).

**Table 2:** Participant characteristics.

|  | Received progesterone |  | p-value* |
| --- | --- | --- | --- |
|  | No<br>(N=1109) | Yes<br>(N=834) |  |
| Suture type received-N (%) |  |  |  |
| Monofilament suture | 553 (50) | 408 (49) | 0.68 |
| Braided suture | 556 (50) | 426 (51) |  |
| Participant characteristics |  |  |  |
| Gestational age at cerclage placement (weeks) |  |  |  |
| Median [IQR, N] | 16.3 [14.1-19.6, 1109] | 15.9 [13.7-19.4, 834] | 0.02 |
| Minimum-Maximum | 8.3-25.7 | 5.1-28.6 |  |
| Maternal age at cerclage placement (years) |  |  |  |
| Mean (SD, N) | 32.7 (4.9, 1109) | 33.2 (5.1, 834) | 0.06 |
| Minimum-Maximum | 18.2-52.3 | 18.6-48.3 |  |
| Ethnicity-N (%) |  |  |  |
| White | 600 (55) | 486 (59) | 0.01 |
| Asian | 207 (19) | 176 (21) |  |
| Black | 239 (22) | 127 (15) |  |
| Mixed | 39 (3) | 32 (4) |  |
| Other | 14 (1) | 8 (1) |  |
| Missing | 10 | 5 | - |
| Pregnancy history |  |  |  |
| Number of previous live pre-term births-N (%) |  |  |  |
| 0 | 726 (65) | 582 (70) | - |
| 1 | 310 (28) | 206 (25) |  |
| 2 | 61 (6) | 39 (5) |  |
| ≥3 | 12 (1) | 7 (<1) |  |
| Median [IQR, N] | 0 [0-1, 1109] | 0 [0-1, 834] | 0.04 |
| Number of previous mid trimester losses-N (%) |  |  |  |
| 0 | 528 (48) | 360 (43) | - |
| 1 | 391 (35) | 344 (41) |  |
| 2 | 153 (14) | 104 (13) |  |
| ≥3 | 37 (3) | 26 (3) |  |
| Median [IQR, N] | 1 [0-1, 1109] | 1 [0-1, 834] | 0.24 |
| Clinical characteristics |  |  |  |
| Primary indication for cerclage-N (%) |  |  |  |
| Deemed risk of preterm birth through history or ultrasound | 642 (58) | 522 (63) | 0.03 |
| Insertion of cervical sutures in previous pregnancies | 237 (21) | 179 (21) |  |
| History of midtrimester loss/premature birth + shortened cervix | 198 (18) | 120 (14) |  |
| History of ≥3 previous midterm losses/premature births | 32 (3) | 13 (2) |  |
| Cerclage technique include bladder dissection-N (%) | 188 (17) | 285 (34) | <0.01 |
| Missing | 0 | 3 | - |
| Previous cervical surgery-N (%) | 285 (26) | 227 (27) | 0.44 |
| Missing | 2 | 2 | - |

\*p-value derived from univariate tests.

Pregnancy loss occurred in 49 (5.9%) of 832 women who received progesterone and 91 (8.3%) of 1103 women who did not receive progesterone (adjusted risk ratio 0.70, 95% CI: 0.50 to 0.99; adjusted risk difference −0.02, 95% CI: −0.04 to −0.001) as per table 3.

**Table 3:** Pregnancy loss.

|  | Received progesterone |  | Risk Ratio<sup>1</sup><br>(95% CI) | Risk Difference<sup>2</sup><br>(95% CI) |
| --- | --- | --- | --- | --- |
|  | No<br>(N=1109) | Yes<br>(N=834) |  |  |
| Pregnancy loss-N (%) |  |  |  |  |
| Yes | 91 (8.3) | 49 (5.9) | 0.70 (0.50 to 0.99) | -0.02 (-0.04 to -0.001) |
| No | 1012 (91.8) | 783 (94.1) |  |  |
| Missing | 6 | 2 |  |  |
| Type of pregnancy loss-N (%) |  |  |  |  |
| Miscarriage | 48 (4) | 22 (3) | - | - |
| Spontaneous | 45 | 20 |  |  |
| Missed | 1 | 1 |  |  |
| Septic | 2 | 1 |  |  |
| Termination | 16 (1) | 4 (<1) |  |  |
| Fetal anomaly | 6 | 1 |  |  |
| Maternal medical condition | 1 | 0 |  |  |
| Maternal sepsis | 7 | 1 |  |  |
| Other<sup>3</sup> | 2 | 2 |  |  |
| Stillbirth | 8 (1) | 10 (1) |  |  |
| Other<sup>4</sup> | 1 (<1) | 0 (-) |  |  |
| Neonatal death <7 days | 18 (2) | 13 (2) |  |  |

<sup>1</sup>Adjusted for primary indication for cerclage, number of previous mid trimester losses, number of previous pre-term births (<34 weeks), previous cervical surgery, cerclage technique involved bladder dissection, suture type received and maternal age at cerclage placement (ethnicity removed from the model due to modelling issues). Values <1 favour received progesterone.
<sup>2</sup>Adjusted for primary indication for cerclage, number of previous mid trimester losses, number of previous pre-term births (<34 weeks), previous cervical surgery, cerclage technique involved bladder dissection and suture type received (ethnicity removed from the model due to modelling issues and maternal age at cerclage placement removed from the model due to convergence issues). Values <0 favour progesterone.
<sup>3</sup>Other includes termination due to SRM and increased risk of sepsis.
<sup>4</sup>Other includes a delivery <20 weeks, unknown if baby was born alive and subsequently died (neonatal death <7 days) or died prior to delivery (miscarriage).

For the key secondary analysis of time from conception to pregnancy end, women who received progesterone had a higher incidence of live birth (sub-distribution hazard ratio 1.12, 95% CI: 1.02 to 1.23). However, these live births appeared to occur at a marginally faster rate (cause specific hazard ratio 1.07, 95% CI: 0.97 to 1.18) (table 4, figure 1). There were no observed differences between the treatment groups regarding the other maternal and neonatal secondary outcomes as reported in table 4. There was no evidence of varying effects in the prespecified subgroup analyses (table 5).

**Figure 1:**
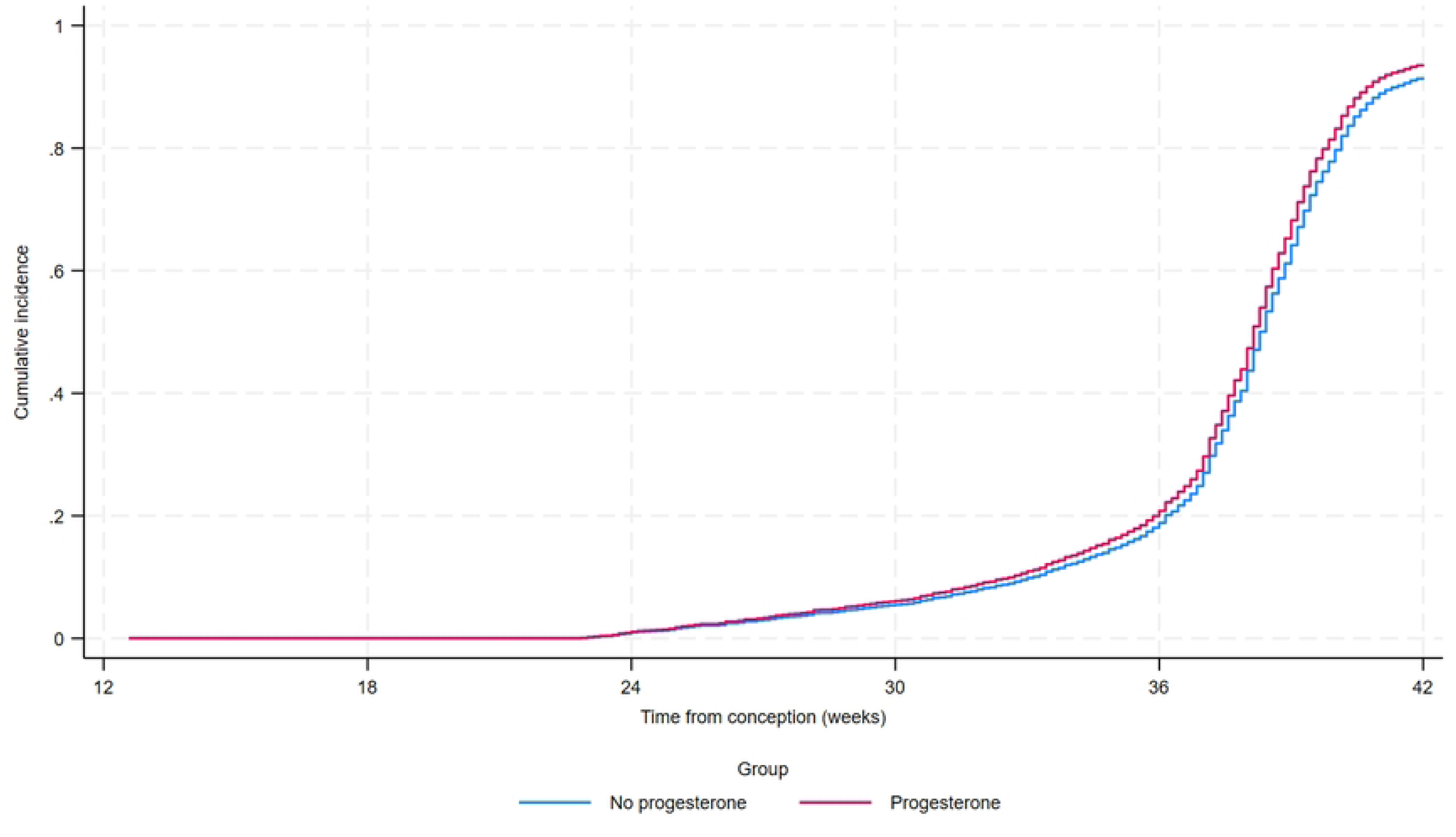
Cumulative incidence function for time to live birth (surviving at least 7 days)

**Table 4:**
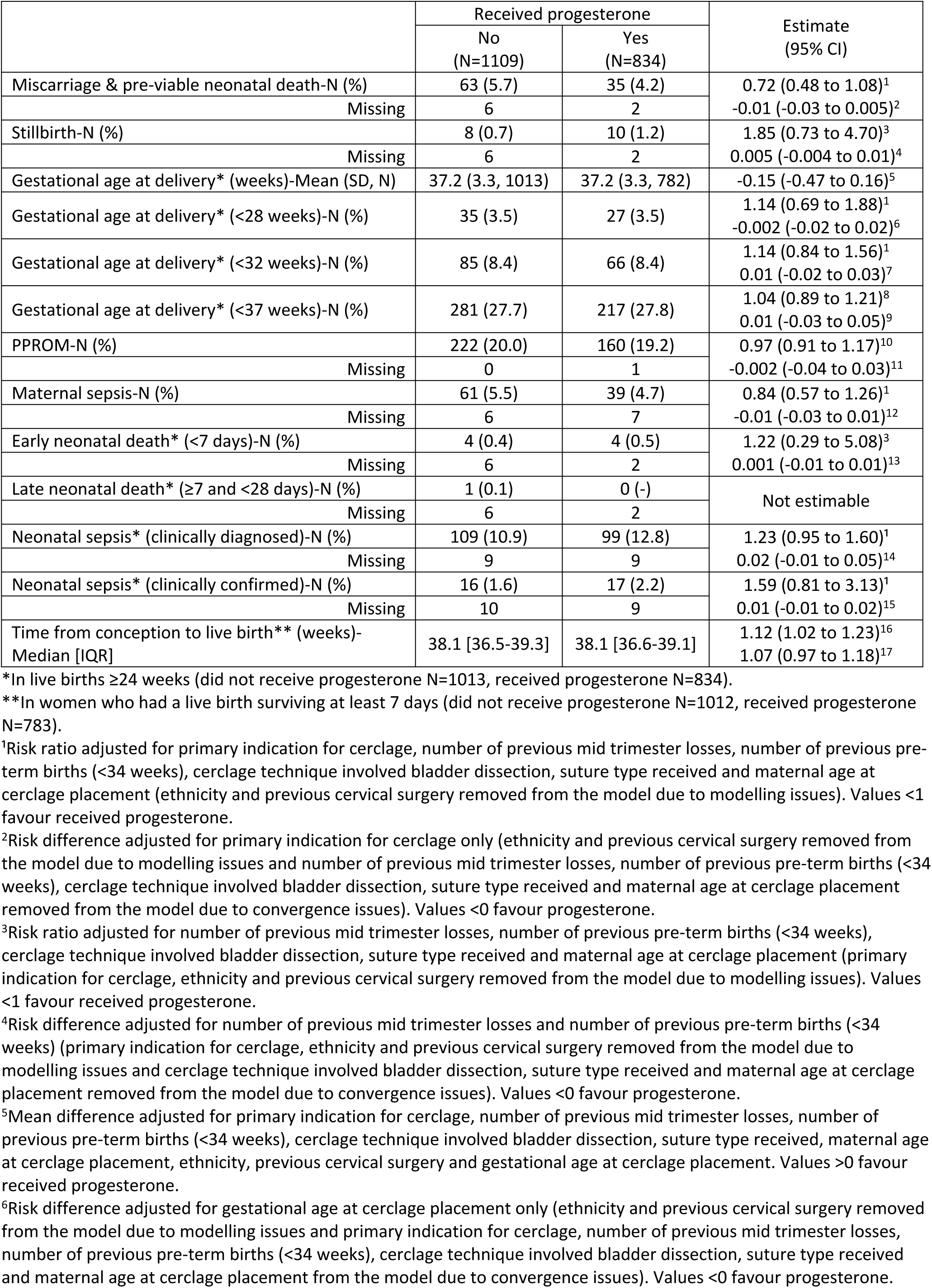
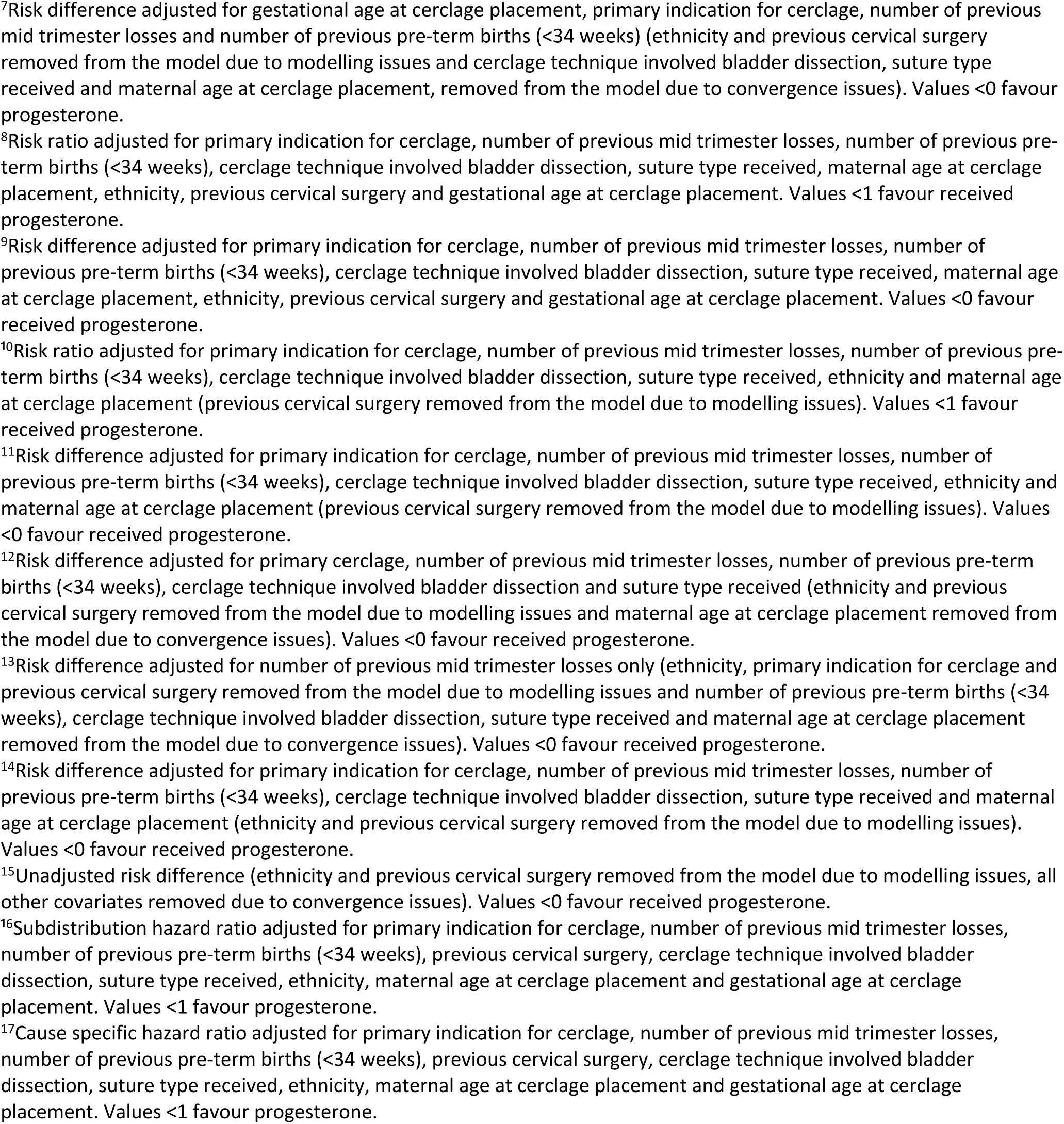
Secondary maternal and neonatal outcomes.

**Table 5:**
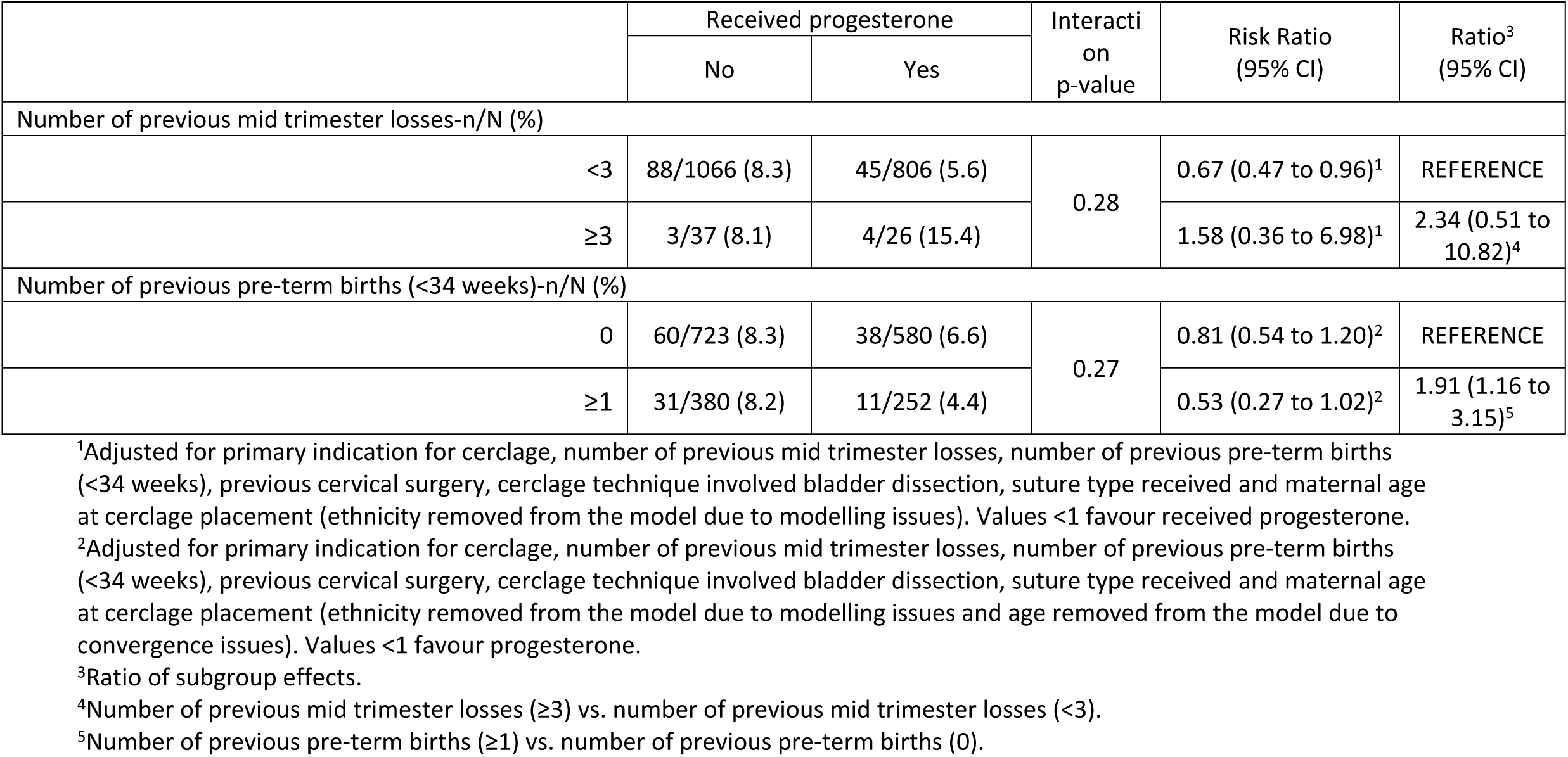
Subgroup analyses for the primary outcome.

## Comment

### Principle findings

In women undergoing a vaginal cervical cerclage due to an increased risk of preterm birth, the addition of progesterone was associated with a 30% relative reduction in pregnancy loss. This should be interpreted taking into account the uncertainty within the estimates and thus the reduction could be from 1-50%.

### Results in the context of what is known

The finding that progesterone potentially reduces pregnancy loss from 8.3% to 5.9% is in keeping with the relative risk reduction seen with vaginal progesterone in the individual patient meta-analysis within EPPIC.(2) The effect size seen within our results is supported by the EPPIC data. These results support the hypothesis that the use of vaginal progesterone therapy in addition to vaginal cervical cerclage is associated with a reduction in pregnancy loss and therefore this may be an important advancement in the prevention of prematurity.

### Clinical implications

Prematurity is the leading cause of perinatal and all-cause mortality for children under 5 years.(11) Therefore while research needs to identify new ways to identify and treat those at risk, it is also important to maximise the benefit from existing therapies and identify in which women these therapies are effective. We highlight an important combination of interventions, which together appear to provide benefit.

### Research implications

To address the limitations of this study and the potential unknown confounders within CSTICH it would be important to consider conducting a randomised controlled trial of progesterone and cervical cerclage as a combination therapy.

### Strengths and limitations

CSTICH was a large UK based randomised controlled trial with recruitment across 75 maternity units. The results presented in this manuscript are from a secondary analysis of the women within the trial and include a large number of women. The strengths of the cohort are thus the size of the cohort, the generalisability to the population of women in the UK receiving a vaginal cervical cerclage and the number of potential confounders and the robustness of the data that was collected. Limitations are the pragmatic nature of the trial and the possibility of unknown confounders that we could not account for in the statistical analysis. For example, there was variation in how progesterone was utilised across the sites. Most women commenced combination treatment around the time of cerclage placement suggesting that the history, clinical findings +/- the results of an ultrasound scan supported the decision for combined therapy to prevent pregnancy loss and prematurity. Approximately 25% of the cohort started progesterone treatment before the placement of a cervical cerclage suggesting a further change in clinical condition perhaps through continued shortening of the cervical length resulted in the placement of a cerclage and 39% of women commenced progesterone after placement of the cervical cerclage for presumed continued concern regarding the risk of pregnancy loss and prematurity. It could be extrapolated that the group who received combined treatment were considered to be at higher risk of a poor outcome but despite this combination treatment improved pregnancy loss rates overall.

There are also known variations within the cervical cerclage technique as reported in the trial results paper. Clinicians were able to determine the type of vaginal cervical cerclage that was inserted (high with bladder dissection or low cervical) based on history, ultrasound or examination findings or personal preference. Women who received progesterone were more likely to have a high vaginal cerclage involving bladder dissection. Our analysis was adjusted for cerclage technique but it should be recognised that this is likely to reflect variation within care recommendations across the preterm birth network with some sites offering as standard a high vaginal cerclage and combination treatment with progesterone. This highlights the importance of optimising outcomes in women undergoing a cervical cerclage to ensure that pregnancy loss is minimised and using outcomes in this high-risk group to determine the best standard operative practices and outcomes.

### Conclusions

The C-STICH cohort represents the largest data set of women receiving cerclage and progesterone to date. In the C-STICH cohort, the addition of vaginal progesterone therapy for women undergoing vaginal cervical cerclage was associated with a 30% relative reduction in the risk of pregnancy loss. There is a need for a randomised controlled trial to determine the women most likely to benefit, the timing and dosage of progesterone therapy and the true effect size.

## Data Availability

The data underlying the results presented in the study are available from The University of Birmingham clinical trials unit by contacting the lead author as per the correspondence details.

